# Shared trans-ancestry architecture of HLA-mediated disease risk in the *All of Us* Research Program

**DOI:** 10.64898/2026.06.26.26356709

**Authors:** Kwangmi Ahn, John S. House, Adam Burkholder, Tam C. Tran, Joseph H. Breeyear, Cristina M. Justice, Jacqueline Durney, Alyssa M. Jones, Parker S. Reyes, Matthew H. Bailey, Mary F. Davis, Anthony T. Vicenti, Jason H. Karnes, Jill A. Hollenbach, David C. Fargo, Geoffrey S. Ginsburg, Richard P. Woychik, Josh C. Denny, Alison A. Motsinger-Reif

## Abstract

The human leukocyte antigen (HLA) region is the strongest genetic contributor to many immune-mediated diseases, yet whether HLA architecture is shared across ancestries remains unclear. We analyzed high-resolution HLA variation in 390,823 participants from the *All of Us* Research Program spanning six genetic ancestry groups, including 262,915 with linked electronic health records. Using whole-genome sequencing and graph-based inference, we genotyped 20 HLA genes at G-group resolution and identified 4,780 distinct alleles. Analyses accounting for disparate sample sizes demonstrated that ancestry-private allelic variation reflected unequal discovery depth rather than ancestry-population specificity. A meta-analysis of ancestry-stratified phenome-wide association analyses with 363 HLA alleles with frequency > 0.001 and 3,430 clinical phenotypes identified 1,461 significant HLA-phenotype associations (FDR < 0.05). Although many associations reached significance in only one ancestry group, effect directions were largely concordant, highlighting differences in allele frequency, linkage disequilibrium, and statistical power among ancestry groups. Stepwise conditional modeling demonstrated that common complex trait variation could be concurrently explained by five to seven independent HLA allele signals. These findings demonstrate that a multi-ancestry, phenome-wide study can distinguish true biological heterogeneity from sampling-driven detectability differences in HLA.

## INTRODUCTION

Genetic variation in the human leukocyte antigen (HLA) region has been robustly associated with numerous autoimmune and inflammatory diseases, infectious disease susceptibility, transplant outcomes, and adverse drug reactions^1–4^. The HLA region on chromosome 6 encodes major histocompatibility complex (MHC) proteins and represents one of the most highly polymorphic regions of the human genome, playing a central role in adaptive immunity^5–7^. HLA molecules present peptide antigens to T cells and thereby shape immune recognition of pathogens, self-antigens, and environmental exposures^8^. Landmark studies have established HLA variation as the strongest genetic contributor to diseases such as type 1 diabetes, rheumatoid arthritis, multiple sclerosis, and celiac disease^9–12^. Despite these advances, we still lack a population-scale understanding of how HLA variation contributes to disease risk across the phenome and whether these associations reflect shared biology across ancestries or ancestry-specific architectures. Further, the focus of most large-scale genetic studies of HLA on individuals of European ancestry^13,14^ may have mischaracterized the global landscape of HLA-mediated disease risk.

Advances in whole-genome sequencing (WGS) and computational HLA inference have enabled high-resolution (G-group) genotyping in population-scale cohorts without reliance on clinically ascertained registries^15–17^. Large biobanks link these data to longitudinal electronic health records (EHR), enabling phenome-wide association studies (PheWAS) across thousands of clinical phenotypes^18,19^. Recent multi-ancestry sequencing studies have revealed substantial differences in allele frequency distributions and uncovered previously unrecognized HLA variation and disease associations^20–22^. However, multi-ancestry HLA-focused phenome-wide analyses remain limited, leaving cross-ancestry generalizability unresolved.

The *All of Us* Research Program prioritizes the inclusion of populations historically underrepresented in biomedical research and integrates WGS with harmonized longitudinal EHR data at a national scale^23–25^. Because HLA is highly polymorphic and structured by ancestry, apparent population-specific variation and association signals may arise from differences in sampling depth, allele frequency, and linkage disequilibrium (LD) as much as from distinct biology. Using the multi-ancestry data available in *All of Us*, we conducted HLA genotyping and PheWAS analyses to evaluate whether HLA disease architecture is truly population-specific or instead reflects shared biology that is differentially detectable across ancestries.

Our work in *All of Us* addresses three important questions: 1) how is HLA diversity distributed across ancestry groups after accounting for disparate discovery depth?; 2) are HLA-phenotype associations identified through phenome-wide analysis predominantly shared or are they ancestry-enriched across populations?; and 3) do dense allele-level association signals resolve into a smaller number of independent effects with conditional modeling? By integrating HLA diversity, cross-ancestry association patterns, and conditional signal resolution, we provide a framework for understanding how HLA-mediated disease risk is shared, structured, and detected across populations. Further, we enable the complex interrogation, visualization, and download of results through the HLA PheWAS Explorer, an extensive HLA dashboard.

## METHODS

### Study participants

The *All of Us* Research Program is a large, longitudinal cohort that enrolls a diverse population across the United States^23–25^. Details of recruitment, consent, and core data collection are described elsewhere.^24^ *All of Us* collects physical measurements, survey data, longitudinal electronic health records (EHR), and genomic information. The current Curated Data Repository release (CDR v8) includes 633,540 participants, including 414,828 with available short-read whole-genome sequencing (srWGS) data and 393,596 with linked EHR data. EHR data were contributed by more than 50 healthcare provider organizations, and participant portal linkages were harmonized using the Observational Medical Outcomes Partnership (OMOP) Common Data Model.^26^

All participants provided informed consent, and the majority (>90%) authorized research use of EHR data under HIPAA, including return of genomic results. The *All of Us* protocol was approved by the central Institutional Review Board (2021-02-TN-001). Genetic ancestry was assigned by projection onto principal components derived from a harmonized reference panel combining the Human Genome Diversity Project and the 1000 Genomes Project (N = 3,942)^27^. Participants were assigned to six continental genetic ancestry groups based on genome-wide genetic ancestry inference: African (AFR), Admixed American (AMR), East Asian (EAS), European (EUR), Middle Eastern (MID), and South Asian (SAS). To minimize confounding from relatedness, we excluded one individual from each pair with estimated identity-by-descent (IBD) > 0.2. We further restricted analyses to participants with sex at birth recorded as male or female, consistent with covariate modeling, and excluded quality-flagged samples prior to analysis.

HLA diversity and LD analyses were conducted in the full set of unrelated, quality-passed individuals with available srWGS data prior to restriction to those with linked EHR data, to maximize representativeness and avoid ascertainment bias arising from differential EHR enrollment across ancestry groups. Because EHR linkage rates varied by ancestry group, restricting analyses to the EHR-linked sample would have introduced differential sampling of HLA diversity; diversity and LD results thus reflect the larger, pre-linkage cohort. PheWAS were restricted to participants with linked EHR data and sex at birth recorded as male or female.

### HLA genotyping and quality control

HLA alleles were inferred from srWGS data using Kourami (v0.9.6) ^28^ with reference to the IPD-IMGT/HLA database (v3.47.0). Kourami provides high-resolution HLA genotyping at G-group resolution (i.e., grouping alleles with identical protein sequences in the antigen recognition domain). A total of 20 HLA loci were analyzed, including classical class I genes (*HLA-A, HLA-B, HLA-C*), classical class II genes (*HLA-DRA, HLA-DRB1, HLA-DRB3, HLA-DRB5, HLA-DQA1, HLA-DQB1, HLA-DPA1, HLA-DPB1*), and non-classical genes (*HLA-DMA, HLA-DMB, HLA-DOA, HLA-DOB, HLA-F, HLA-G, HLA-H, HLA-J, HLA-L*). Because *HLA-DOB* was not included in the default Kourami locus list, we modified the source code to incorporate this gene.

Processing was performed within the *All of Us* Researcher Workbench using a containerized GATK environment (us.gcr.io/broad-gatk/gatk:4.3.0.0). The alignAndExtract_hs38DH.sh script was modified to retrieve unmapped reads and reads mapping to HLA loci directly from cloud-hosted CRAM files using GATK PrintReads. In rare instances of failed assembly at *HLA-DOA*, the locus was excluded, and the pipeline was rerun. Alleles were reported at G-group resolution (G-groups comprise alleles sharing identical nucleotide sequences in exons 2 and 3 for class I loci, and exon 2 for class II loci).

Genotype calls were excluded if they were ambiguous, if they showed <95% sequence identity to the reference sequence, or if they had a MaxFlow score <10.

### HLA diversity and down-sampling analyses

We calculated allele frequencies separately within each continental ancestry group (AFR, AMR, EAS, EUR, MID, and SAS) using the pre-EHR-linkage unrelated sample of 390,823 individuals. We quantified the number of alleles observed per locus, as well as alleles unique to a single ancestry group and alleles shared across groups. We also evaluated the number of alleles required to account for 90% of cumulative allele frequency per locus and ancestry group as a measure of effective diversity that is robust to ultra-rare variants. Expected heterozygosity (H = 1 − Σpᵢ²) was computed as an additional summary measure of allelic diversity.

Because the European ancestry group was substantially larger than other groups, we implemented a downsampling framework to determine the contribution of sample size to apparent allele discovery. We generated 20 independent subsets of 50,000 European ancestry individuals using sex-stratified random sampling from the EHR-linked analytic cohort while preserving the observed male/female ratio. For each replicate, we recalculated the numbers of observed, private, and shared alleles. We used mean values across replicates to estimate sampling variability and evaluate whether apparent ancestry-private alleles reflected differences in statistical power rather than true population specificity.

### Linkage disequilibrium analysis

We evaluated linkage disequilibrium (LD) among 20 classical and non-classical HLA loci within the AFR, AMR, and EUR ancestry groups. Corresponding matrices for EAS, SAS, and MID are provided in Supplementary Data 1. We calculated LD at the allele level using the squared Pearson correlation coefficient (r²) between allele dosage vectors at G-group resolution, restricting analyses to individuals with non-missing genotype calls at both loci. LD was assessed only between alleles at distinct loci, as within-locus correlations in multi-allelic systems are structurally constrained. Sample sizes for LD analyses reflect the pre-EHR-linkage unrelated cohort as described in the Study participants section.

### Phenome-wide association studies

We conducted PheWAS in the three largest ancestry groups (EUR, AFR, and AMR) using the PheTK package (v0.2.1rc5)^29^. ICD-9 and ICD-10 codes were mapped to 3,430 phecodes using PhecodeX v1.0^30^. A phecode was defined as a case if the corresponding diagnosis occurred on at least two distinct dates in the EHR. Phecodes with fewer than 50 cases or 50 controls within an ancestry group were excluded.

Within each ancestry group, we required HLA alleles to have a minimum frequency of 0.001. This threshold leveraged the large sample size while retaining adequate statistical power. To ensure stable model estimation, we further restricted analyses to allele-phecode pairs with at least 50 carriers and 50 non-carriers. We tested associations under a dominant genetic model using logistic regression, adjusting for age at last EHR encounter, sex at birth, duration of EHR follow-up (defined as the time between first and last recorded EHR encounters), and the first 15 genetic principal components derived within each ancestry group.

To assess robustness to genetic model specification, we repeated analyses under an additive genetic model, coding allele dosage as 0, 1, or 2 copies of the allele of interest. We restricted these analyses to the same set of allele-phecode pairs evaluated under the dominant model.

We combined ancestry-specific summary statistics using fixed-effects inverse-variance-weighted meta-analysis implemented in METAL^31^. We assessed between-ancestry heterogeneity using Cochran’s Q statistic and quantified it using the I² metric. We controlled for multiple testing using the Benjamini-Hochberg FDR procedure and considered associations with FDR < 0.05 statistically significant.

### Conditional fine-mapping

To identify independent HLA association signals, we performed forward stepwise conditional analyses for five phenotypes pre-specified based on having the largest number of FDR-significant alleles in the primary meta-analysis, including type 1 diabetes, celiac disease, hypothyroidism, multiple sclerosis, and rheumatoid arthritis. Index alleles for each phenotype were defined as HLA alleles reaching FDR significance (q < 0.05) in the primary cross-ancestry meta-analysis. Conditional modeling was conducted separately within AFR, AMR, and EUR ancestry groups using logistic regression models adjusted for age, sex, EHR length, and ancestry principal components. At each step, remaining index alleles were tested while conditioning on alleles retained in prior steps, and ancestry-specific effect estimates were combined using fixed-effects inverse-variance meta-analysis. Independent signals were defined using a Bonferroni-corrected significance threshold of 0.05/M, where M is the number of index alleles for that phenotype, applied to the meta-analysis p-value at each step. Stepwise selection terminated when no remaining allele exceeded this threshold. To reduce redundancy arising from strong within-gene LD, at most, one allele per HLA gene was retained during stepwise selection. Final joint models were subsequently refit within each ancestry group, including all retained alleles simultaneously, and ancestry-specific estimates were combined using fixed-effects inverse-variance meta-analysis to generate the reported conditional effect estimates and confidence intervals. Linkage disequilibrium among retained alleles was subsequently evaluated to distinguish conditionally independent effects from associations driven by extended haplotypes. Additional implementation details are provided in the Supplementary Methods.

### Novelty classification of HLA–phenotype associations

To identify candidate novel HLA allele–phenotype associations, we applied a structured screening strategy, retaining associations that reached FDR-corrected significance (q < 0.05) in cross-ancestry meta-analyses under a dominant model and showed concordant direction of effect in at least two ancestry groups.

Prior HLA associations were curated from the NHGRI-EBI GWAS Catalog (v1.0, full release)^32^, an HLA-focused PheWAS catalog^33^, a curated HLA association dataset^22^, and a general PheWAS reference catalog^34^. Alleles were harmonized to HLA nomenclature by standardizing formats and resolving representations up to three-field resolution, with comparisons performed at the maximum shared resolution between query and reference alleles. Phenotype labels were normalized for comparison.

Novelty was assigned hierarchically by comparison to phenotypically matched prior records at the highest shared allele resolution. Exact matches were classified as not novel; associations with prior locus-level but not allele-level evidence were classified as gene-known/allele unresolved; associations lacking prior gene-level evidence were classified as gene-level novel; and associations within known genes without matching allele-level evidence were classified as allele-level novel.

To account for HLA region LD, we applied a two-stage LD-aware procedure. Associations in strong LD (r² ≥ 0.8) with previously reported alleles were removed using an allele-level LD graph constructed across ancestry groups. Remaining signals were collapsed into locus-level associations by grouping alleles within known haplotype blocks (e.g., *DRB1*/*DRB3*/*DRB5*) and phenotype categories, retaining the most significant association per locus–phenotype group.

Finally, novelty assignments were refined through a targeted literature review by requiring the absence of prior gene-level evidence and/or matching allele-level reports for classification as novel.

### HLA PheWAS results and HLA calls

We developed an accompanying interactive HLA PheWAS resource to enable transparent exploration of the full association results across alleles, traits, and ancestries. The PheWAS Explorer provides users the opportunity to explore results beyond static summary figures and tables by querying individual HLA genes, alleles, phecodes, and ancestry strata; visualizing association landscapes with Manhattan-style plots, forest plots, and multi-trait heatmaps; and examining both effect estimates and significance metrics in the context of sample size, case and control counts, and allele frequency differences. Additional views highlight ancestry-specific findings, bidirectional allelic effects across phenotypes, and higher-resolution drilldown of allele families, supporting both broad pattern recognition and focused follow-up of specific signals.

The PheWAS Explorer improves the interpretability, reproducibility, and reuse of the results by providing access to a high-dimensional HLA association dataset without the need for custom re-analysis of the primary results files. It offers a practical framework for identifying whether observed signals are concentrated within specific disease domains, whether associations are shared or differ across ancestry groups, and whether related alleles within a locus have convergent or divergent phenotypic profiles. By serving as a companion analytical interface to the manuscript, the PheWAS Explorer expands the utility of the study from a fixed set of reported findings to a broad community resource for hypothesis generation, validation of reported associations, and future comparative analyses of HLA-disease relationships (https://manticore.niehs.nih.gov/AoU_HLA_PheWAS_Explorer).

High-resolution HLA genotype calls are also available to approved researchers through the *All of Us* Researcher Workbench community workspace (https://workbench.researchallofus.org/).

## RESULTS

### HLA diversity across ancestry groups

We genotyped HLA alleles at G-group resolution across 20 loci in 390,823 unrelated individuals from the *All of Us* Research Program. *All of Us* is one of the largest WGS-based HLA resources available at this resolution, enabling detailed characterization of allele frequency spectra, population-specific diversity, and sharing patterns. From an initial 414,828 participants with srWGS data, we excluded 987 quality-flagged samples and 23,018 related individuals, yielding the pre-EHR-linkage analytic sample (**Supplementary Table 1**). We conducted HLA diversity and LD analyses in this larger sample to avoid ancestry-differential ascertainment. EHR linkage rates varied across ancestry groups (EUR 69.8%; AFR 63.6%; AMR 65.0%; EAS 56.8%; SAS 61.0%; MID 71.9%), such that restricting these analyses to EHR-linked participants would have underrepresented certain ancestry groups in the diversity estimates. Restriction to participants with linked EHR data yielded a final PheWAS analytic cohort of 262,915 individuals. The final cohort comprised 160,407 females and 102,508 males and included 156,802 EUR, 49,410 AFR, 46,916 AMR, 5,492 EAS, 3,247 SAS, and 1,048 MID ancestry individuals (**Table 1**).

**Table 1.** Study cohort sample sizes by genetic ancestry and sex for PheWAS. Number of unrelated participants included in analyses after exclusion of related individuals (identity-by-descent > 0.2) and restriction to individuals with linked electronic health record data. Counts are shown by sex and genetically inferred ancestry group. AFR, African; AMR, Admixed American; EAS, East Asian; EUR, European; MID, Middle Eastern; SAS, South Asian.

| Sex | AFR | AMR | EAS | EUR | MID | SAS | Total |
| --- | --- | --- | --- | --- | --- | --- | --- |
| Female | 30,490 | 30,662 | 3,537 | 93,349 | 584 | 1,785 | 160,407 |
| Male | 18,920 | 16,254 | 1,955 | 63,453 | 464 | 1,462 | 102,508 |
| Total | 49,410 | 46,916 | 5,492 | 156,802 | 1,048 | 3,247 | 262,915 |

Across all participants, we identified 4,780 distinct alleles at G-group resolution. Calling rates exceeded 90% across most loci, though lower rates at *HLA-DRB3* and *HLA-DRB5* reflect haplotype-dependent gene presence rather than systematic inference performance differences (**Supplementary Data 1**).

Allelic diversity varied markedly across loci and ancestry groups (**Fig. 1**). The greatest diversity was observed at classical class I loci that are routinely tested in clinical transplantation, particularly *HLA-B*, followed by *HLA-A* and *HLA-C*. Among classical class II loci, substantial diversity was observed at *HLA-DRB1*, *HLA-DQB1*, and *HLA-DPB1*. In contrast, non-classical loci, including *HLA-DMA*, *HLA-DMB*, and *HLA-DRA*, exhibited limited allelic variation. Non-classical loci also showed comparatively limited variation across ancestry groups. In absolute terms, AFR and EUR exhibited the largest numbers of distinct alleles, reflecting both underlying genetic diversity and differences in sample size.

**Figure 1.**
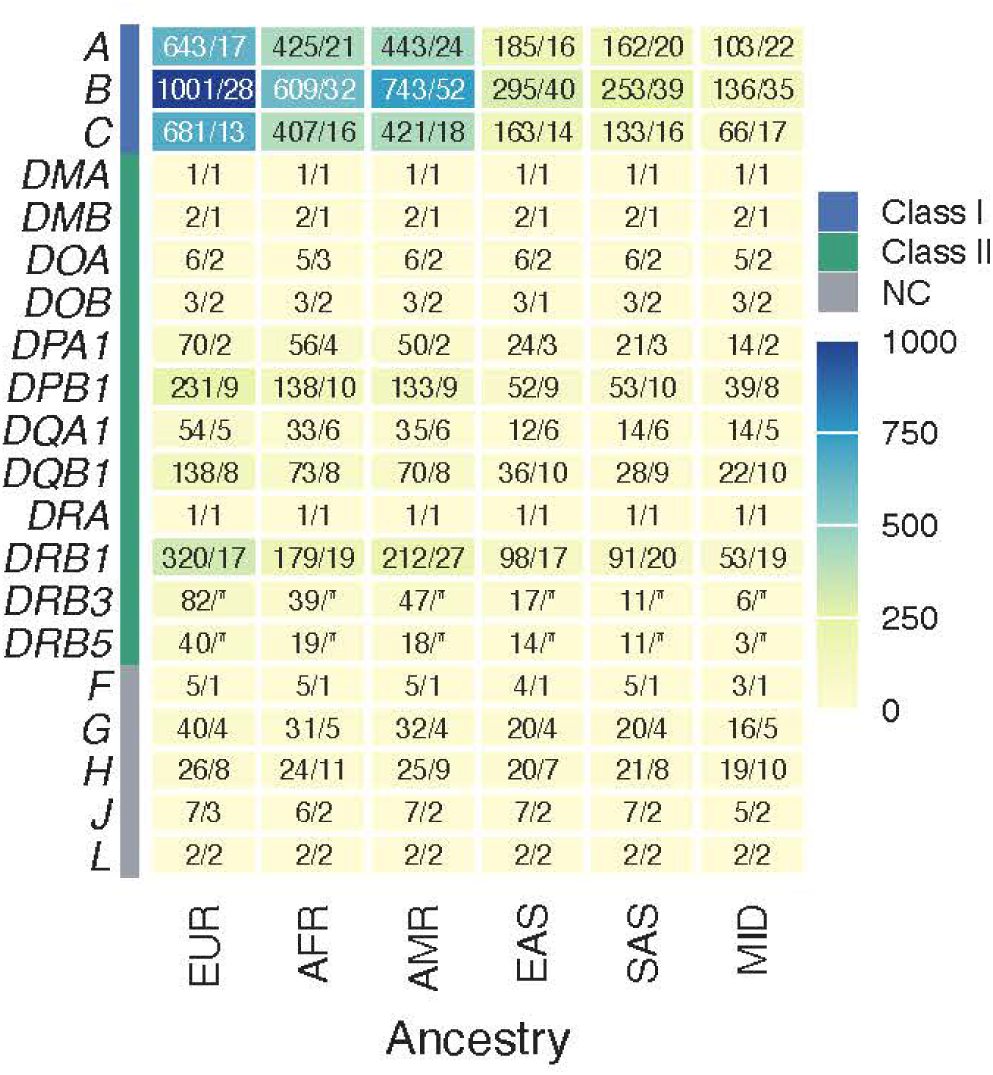
HLA allelic and ancestral diversity. Twenty HLA genes were called with Kourami in *All of Us* participants with srWGS data (v8). Each cell displays the number of alleles called for that ancestry group and the number of alleles needed to call 90% of individuals for that ancestry. Cells are colored based on total number of called alleles per gene. *overall HLA gene call rate < 90%. NC – non-classical.

To quantify effective allelic diversity while reducing sensitivity to ultra-rare variants, we evaluated the number of alleles required to account for 90% of cumulative frequency at each locus (**Fig. 1**). Classical class I and class II loci required substantially more alleles to capture 90% of cumulative frequency than non-classical loci. Among classical class I loci, *HLA-B* required the highest number of alleles across all ancestry groups, with particularly high values in AMR (52 alleles) relative to AFR (32) and EUR (28). Similar but less pronounced patterns were observed at *HLA-A* and *HLA-C*. Among classical class II loci, *HLA-DRB1* showed similar patterns, whereas non-classical loci required fewer alleles to account for cumulative frequency across all groups. These findings demonstrate strong locus-specific differences in effective diversity alongside a broadly shared rare-allele frequency spectrum across populations.

To assess the contribution of sampling depth to apparent ancestry-specific variation, we evaluated allele sharing across the EUR, AFR, and AMR groups (**Fig. 2a**). Of the 4,780 distinct alleles identified across all loci, 3,880 (81.5%) were observed in at least one of the three major ancestry groups. Of these, 1,027 alleles (21.5%) were shared across all three ancestry groups, whereas 2,105 alleles (44.0%) appeared ancestry-private within these three major groups. Down-sampling of EUR to 50,000 individuals (mean of 20 sex-stratified replicates) reduced EUR-private alleles from 1,273 (26.6%) to 596 (12.5%) (**Fig. 2b**), approaching the levels observed in AFR (414 alleles, 8.7%) and AMR (418 alleles, 8.7%). These results demonstrate that a substantial fraction of apparent EUR-private alleles reflects sampling depth rather than strict population specificity.

**Figure 2.**
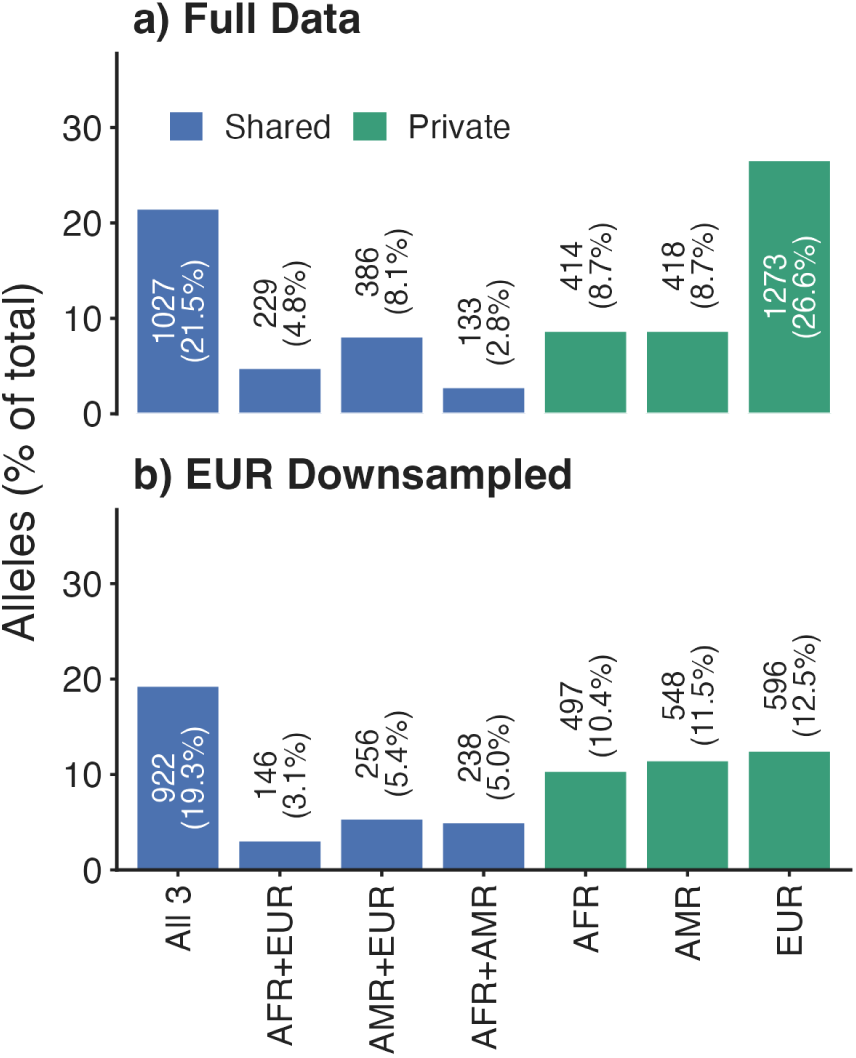
Allele sharing and frequency structure across major ancestry groups. **(a)** Distribution of distinct HLA alleles by shared category across the three major ancestry groups (EUR, AFR, and AMR). Categories include alleles shared across all three groups, shared between exactly two groups, private to a single group. Bars represent the proportion of total distinct alleles; counts are shown above bars. **(b)** Allele sharing after downsampling EUR to 50,000 individuals (mean of 20 sex-stratified replicates). Bars show the mean proportion of alleles in each sharing category following downsampling. Counts displayed above bars represent the mean number of alleles identified.

Expected heterozygosity exhibited similar locus-specific patterns, with the highest diversity at classical loci and less diversity at non-classical loci (**Supplementary Fig. 1**).

### Ancestry-specific linkage disequilibrium architecture across the HLA region

We evaluated allele-level LD across 20 HLA loci within six genetic ancestry groups. Full allele-level LD matrices for all six ancestry groups are provided in **Supplementary Data 2**. The strongest inter-locus coupling localized within the class II region, particularly across the *DRB1–DQA1–DQB1* block. For example, *HLA-DQA1*02:01:01G* showed strong LD with *HLA-DRB1*07:01:01G*, with r² = 0.98 in EUR, 0.96 in AMR, and 0.72 in AFR. Across ancestries, allele-level r² values were generally higher in EUR and AMR relative to AFR, consistent with more extensive extended haplotypes in European-derived populations and greater haplotypic dispersion in individuals with African ancestry.

### Phenome-wide associations in the HLA region across ancestries

Of the 594,316 allele–phecode meta-analysis tests, 1,461 associations met the FDR threshold (q < 0.05) (**Fig. 3**; **Supplementary Data 3**). These associations involved 246 distinct HLA alleles and 455 unique phecodes spanning 17 disease categories.

**Figure 3.**
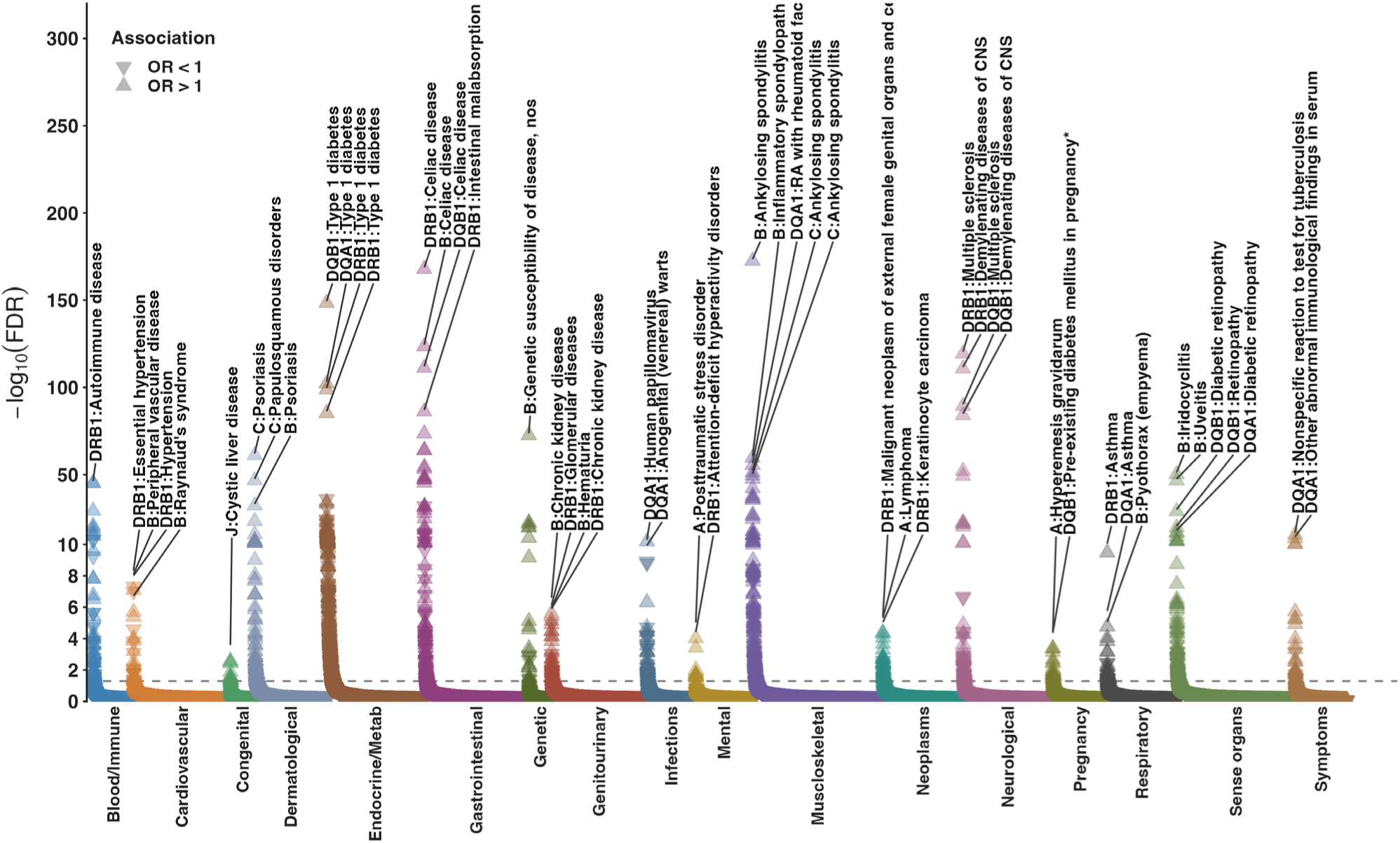
Phenome-wide HLA associations from cross-ancestry meta-analysis. Each point represents an HLA allele–phecode association grouped by phecode category along the x-axis. The y-axis shows −log10(FDR), and the dashed horizontal line indicates the significance threshold corresponding to FDR q = 0.05. Upward and downward triangles denote risk (OR > 1) and protective (OR < 1) associations, respectively. Representative associations are labeled.

Among associations evaluated in at least two ancestry groups (n = 1,229), 80.3% demonstrated concordant effect directions. Between-ancestry heterogeneity was generally limited (median I² = 0%), although 29.3% of associations exhibited I² > 50%. Overall, these findings largely support shared genetic effects across the EUR, AFR, and AMR ancestry groups. Significant associations were enriched within endocrine/metabolic and musculoskeletal phenotypes (**Fig. 4a**), reflecting the broad pleiotropic effects of HLA variation across clinical domains beyond canonical autoimmune and inflammatory diseases. Notably, a subset of associations involved clinical domains not traditionally emphasized in HLA studies, including congenital, cardiovascular, neurological, dermatological, infectious, and reproductive conditions (**Fig. 4a**). While many of these signals occurred in HLA loci with established roles in immune-mediated disease, the corresponding phenotype domains have been less extensively characterized in the context of HLA variation. These findings suggest that phenome-wide approaches may uncover previously underexplored manifestations of HLA-associated risk, although their biological interpretation requires further validation.

**Figure 4.**
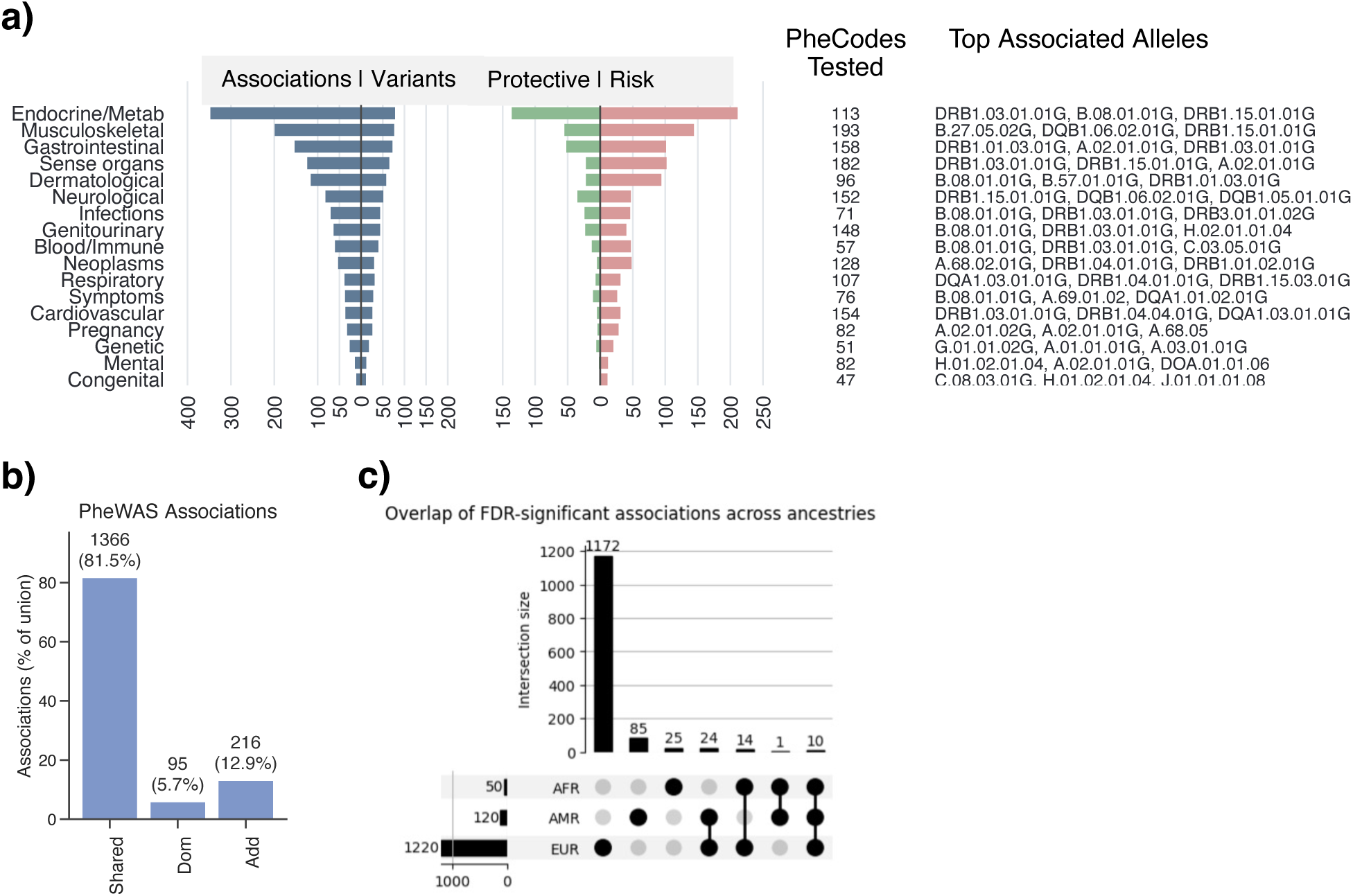
Summary of HLA PheWAS. (a) Distribution of significant HLA–phenotype associations (FDR q < 0.05) across phecode categories from the cross-ancestry meta-analysis. Left panels show the numbers of associations and distinct HLA alleles per category, and right panels show the numbers of protective (OR < 1) and risk (OR > 1) associations. The number of phecodes tested and representative highly associated alleles are shown on the right. (b) Comparison of dominant and additive genetic models for HLA PheWAS. Bars show the numbers of associations shared between models or uniquely identified under dominant or additive encoding. (c) Overlap of FDR-significant HLA–phenotype associations across EUR, AFR, and AMR ancestry groups. Vertical bars indicate the number of shared or ancestry-specific associations, and connected dots indicate ancestry combinations contributing to each overlap set.

To evaluate robustness to genetic model specification, we performed parallel analyses using an additive genetic model. Effect-size estimates were highly correlated among FDR-significant associations (Pearson’s r = 0.997), indicating near-identical direction and magnitude of effects across models. Of the 1,461 associations significant under the dominant model, 1,366 (93.5%) were also significant under the additive model. The remaining 95 were uniquely detected under the dominant model, while 216 additional signals were identified exclusively under the additive model (**Fig. 4b**), yielding 1,582 total associations under the additive model. Dominant-only and additive-only signals exhibited similar allele frequency distributions (Mann–Whitney U test, *P* = 0.16), indicating that differences between models were not driven by allele frequency.

### Ancestry-stratified PheWAS in the HLA region

Within ancestry-specific analyses, 1,220 associations met FDR < 0.05 in EUR, compared to 50 in AFR and 120 in AMR (**Fig. 4c**). Of the 1,282 associations significant in only a single ancestry, 95.9% demonstrated concordant direction of effect in at least one additional ancestry group, although they did not reach FDR significance outside the discovery group. The 85 and 25 associations unique to AMR and AFR populations, respectively, can be seen in the accompanying PheWAS Explorer (https://manticore.niehs.nih.gov/AoU_HLA_PheWAS_Explorer).

Although many associations achieved significance in only one ancestry group, cross-ancestry agreement was stronger for effect direction than for effect magnitude. Among associations evaluated across ancestry pairs (based on the entry set of all FDR < 0.05 associations in any ancestry), 74-83% showed concordant effect signs, with moderate rank-based agreement of effect estimates but weaker linear correlations.

To characterize these ancestry-restricted signals, we examined associations reaching FDR significance in AFR only, AMR only, or in both AFR and AMR but not EUR, restricting the analysis to allele–phecode pairs evaluable across all three ancestry groups. Exemplars are shown in **Fig. 5**, and the complete set is provided in **Supplementary Data 4**. AFR-unique associations included *HLA-DRB*01:02:01G* with lichen planus, *HLA-B*41:02:01G* with calcium deposits in tendon and bursa, and *HLA-DQB1*02:01:01G* with decreased risk of sarcoidosis. AMR-unique associations included *HLA-DRB1*13:01:01G* with pneumonia due to fungus and *HLA*A:02.644* with Graves’ disease. A single association was significant in both AFR and AMR but not in EUR (*DRB1:04:05.01G* and autoimmune disease).

**Figure 5.**
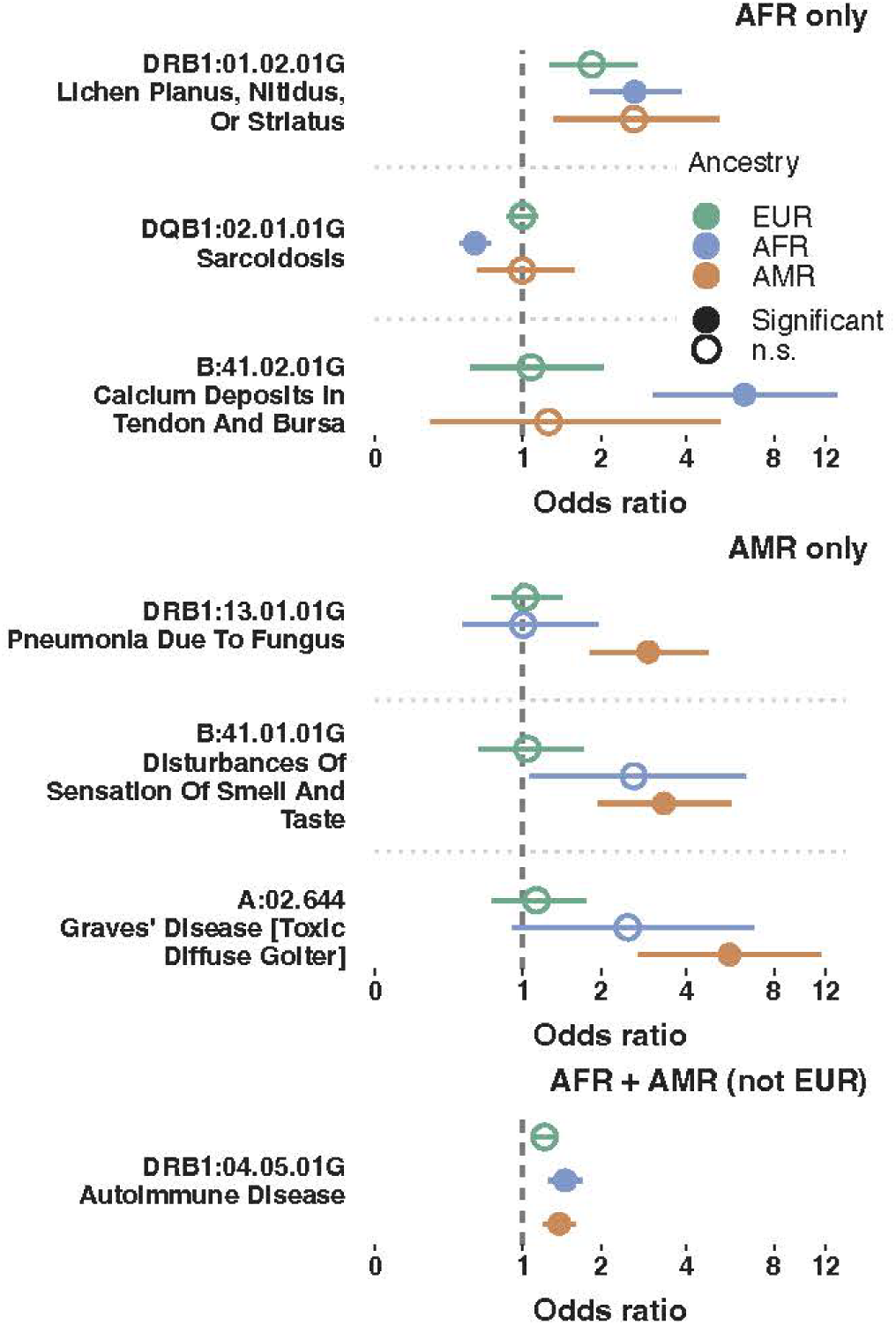
Ancestry-specific associations. Odds ratios and 95% confidence intervals are shown for ancestry-specific exemplars. Solid circle represents FDR q < 0.05 in ancestry specific PheWAS.

### Conditional fine-mapping of HLA associations

Single-variant meta-analyses identified substantial numbers of HLA alleles associated with each phenotype prior to conditional modeling. At FDR < 0.05, 47 distinct alleles were associated with type 1 diabetes, 43 with celiac disease, 31 with hypothyroidism, 25 with multiple sclerosis, and 23 with rheumatoid arthritis.

Stepwise conditional meta-analyses markedly reduced these correlated signals to a limited number of independent gene-distinct or haplotype-distinct effects. Type 1 diabetes resolved into seven independent signals spanning both class II and class I loci, with the strongest conditional effect at *DQB1*03:02:01G* (OR = 3.04). Celiac disease resolved into six independent signals dominated by DQA1 and DRB1 loci. Multiple sclerosis resolved into five gene-distinct effects led by *DRB1*15:01:01G* (OR = 2.88). Rheumatoid arthritis resolved into one risk and four protective alleles in five HLA genes: *HLA-B, HLA-C, HLA-DRB1, HLA-DQA1,* and *HLA-DQB1*. (**Fig. 6**).

**Figure 6.**
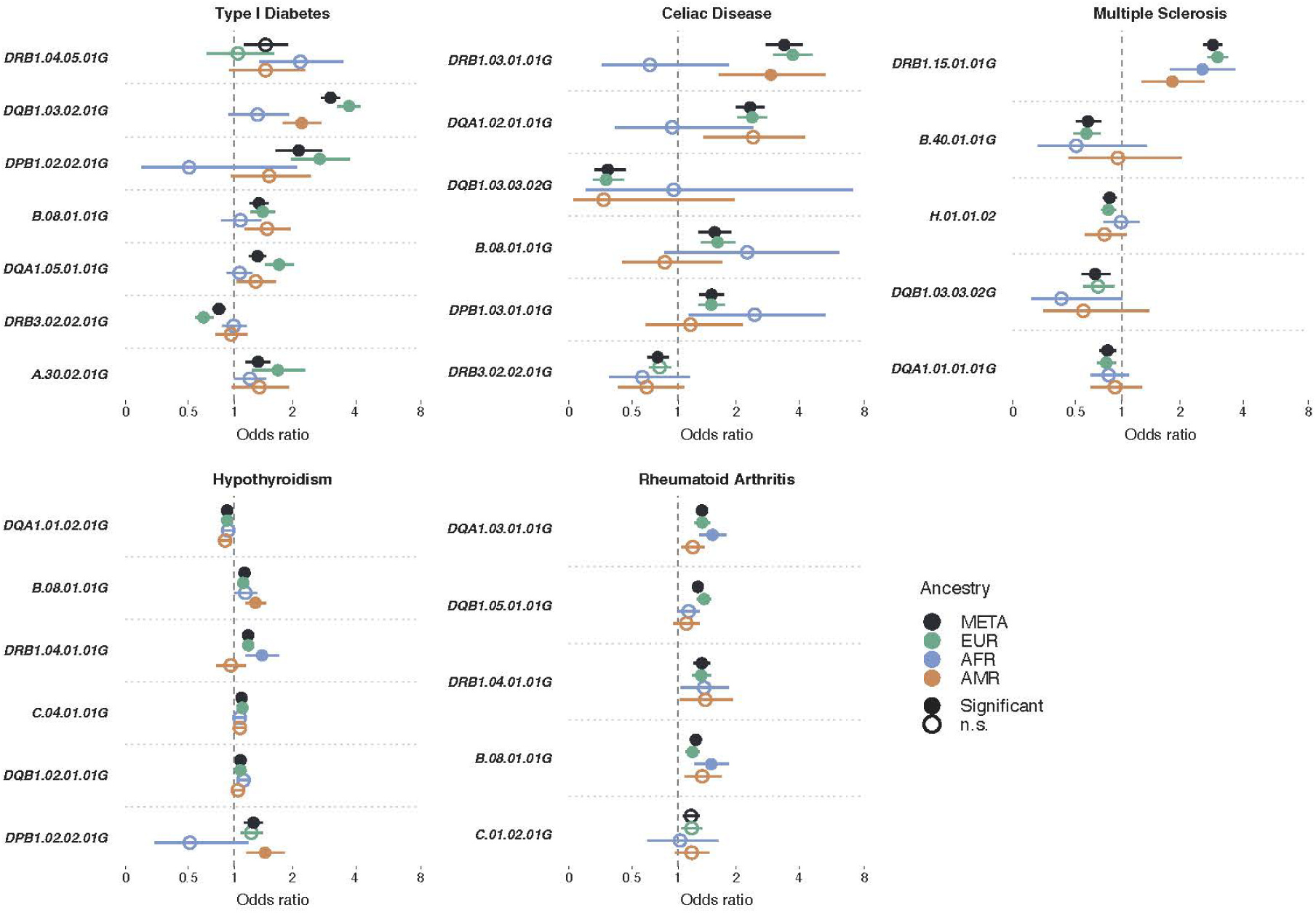
Conditional fine-mapping. Forest plots show conditional odds ratios for independent HLA alleles identified by stepwise joint modeling for five phenotypes. Points show ancestry-specific (EUR, AFR, AMR) and meta-analysis (META) estimates; horizontal lines denote 95% confidence intervals. Filled markers indicate Bonferroni-significant effects.

Across phenotypes, conditional modeling consistently reduced clusters of correlated alleles to five to seven independent gene-level signals, primarily localized to class II loci with additional class I contributions. Independent signals were detected across EUR, AFR, and AMR populations, indicating broadly shared genetic effects despite ancestry-specific LD differences (**Supplementary Fig. 2**). These results demonstrate a recurring architectural pattern in which diverse HLA-associated phenotypes are driven by a compact set of gene-distinct effects embedded within extended MHC haplotypes.

### HLA Pleiotropy

Many HLA alleles were associated with numerous traits across multiple phecode categories (**Fig. 7**). Among the 246 alleles implicated in meta-significant associations, 174 (70.7%) were associated with two or more distinct phecodes at FDR < 0.05, 131 (53.3%) with three or more phecodes, and 81 (32.9%) with five or more phecodes. The most pleiotropic allele, *HLA-DRB1*03:01:01G*, was associated with 61 phecodes spanning 13 disease categories (**Fig. 7**). Pleiotropic signals were concentrated within class II loci and commonly involved combinations of autoimmune, inflammatory, and infectious phenotypes, consistent with shared antigen-presentation pathways and extended haplotypic structure within the MHC. These pleiotropy analyses were based on marginal single-variant associations; the conditional modeling applied to exemplar phenotypes was not extended systematically across all phecode pairs.

**Figure 7.**
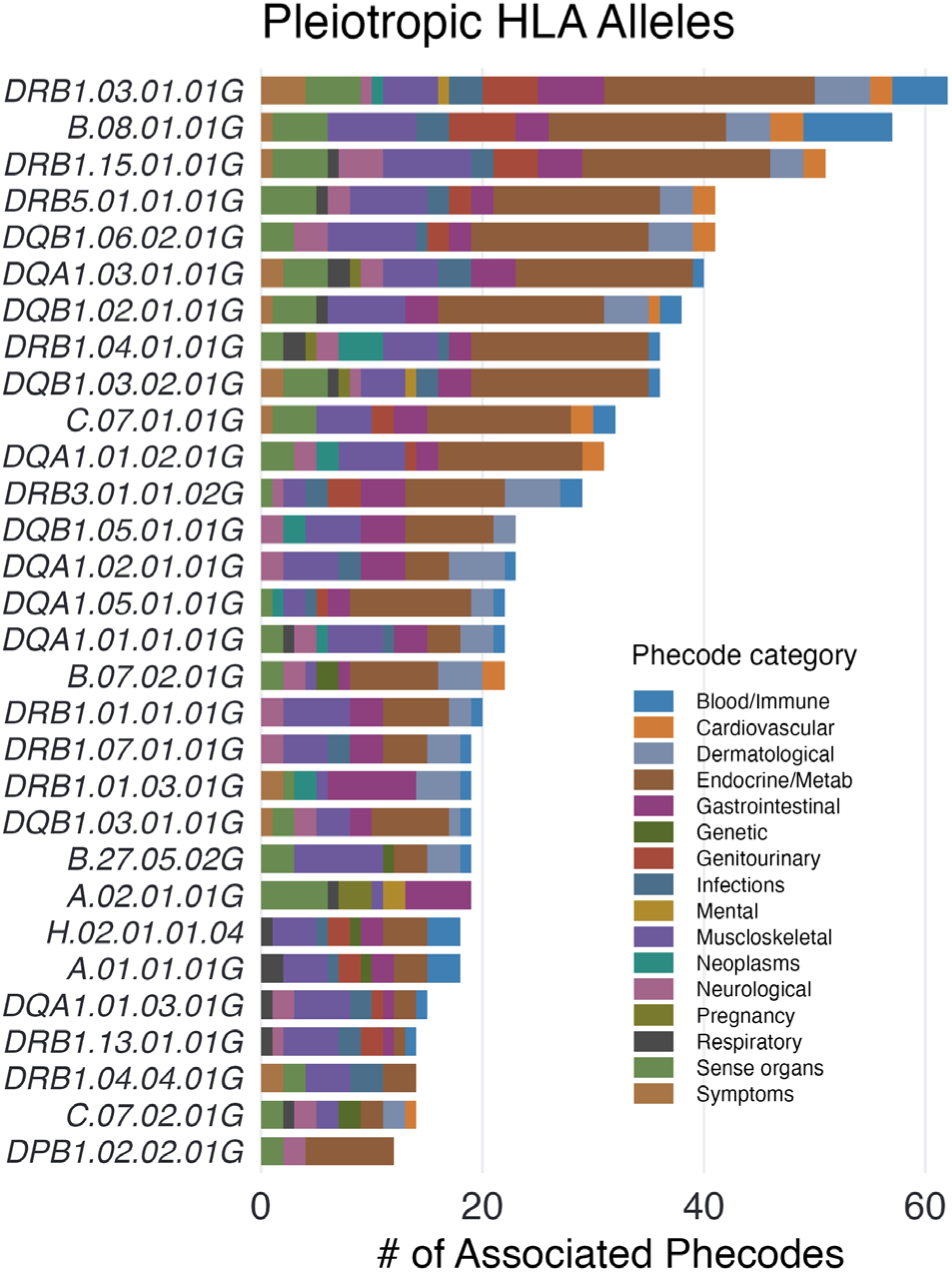
Pleiotropic HLA associations across phenotypes. Stacked bar plot showing the number and distribution of phecode associations (meta-analysis FDR < 0.05) for the top 30 HLA alleles under the dominant model. Colors indicate phecode categories, and bar length reflects the total number of associated phenotypes per allele.

### Novel findings

Of 1,461 HLA-phenotype associations reaching FDR < 0.05, 987 showed concordant directions of effect in at least two ancestry groups. After excluding 653 previously reported associations and removing 273 signals in strong LD (r² ≥ 0.8) with known loci, 61 candidate novel associations remained. LD-aware collapsing within the HLA region yielded 42 independent high-confidence signals.

Following literature curation, all 42 associations were novel at either the gene or allele level, including 16 (38%) candidate gene-level associations and 26 (62%) allele-level refinements. While many signals mapped to classical HLA loci and known immune pathways, gene-level candidates were enriched in non-classical loci (*HLA-DOA*, *HLA-DOB*, *HLA-H*, and *HLA-J*). Notably, several associations extended beyond traditional HLA-linked phenotypes, including infectious, psychiatric, congenital, and structural traits, suggesting a broader role for HLA variation across diverse disease domains (**Supplementary Data 5**).

We highlight two examples with cross-ancestry support. *HLA-DOA*01:02* was associated with Streptococcus pneumoniae infection (OR = 6.18, 95% CI 2.63–14.54, *P* = 3.0 × 10⁻⁵), with concordant effects in AFR (OR = 6.30, *P* = 0.01) and AMR (OR = 6.11, *P* = 8.8 × 10⁻⁴). *HLA-C*15:05* was associated with drug-induced psychotic disorder (OR = 2.44, 95% CI 1.63–3.65, *P* = 1.35 × 10⁻⁵), with consistent effects in EUR (OR = 2.96, *P* = 0.034) and AFR (OR = 2.41, *P* = 1.9 × 10⁻⁴) (**Fig. 8**). These findings demonstrate that LD-aware, cross-ancestry HLA PheWAS identify both allelic refinements within classical loci and candidate gene-level associations at non-classical genes, extending HLA associations beyond canonical immune phenotypes.

**Figure 8.**
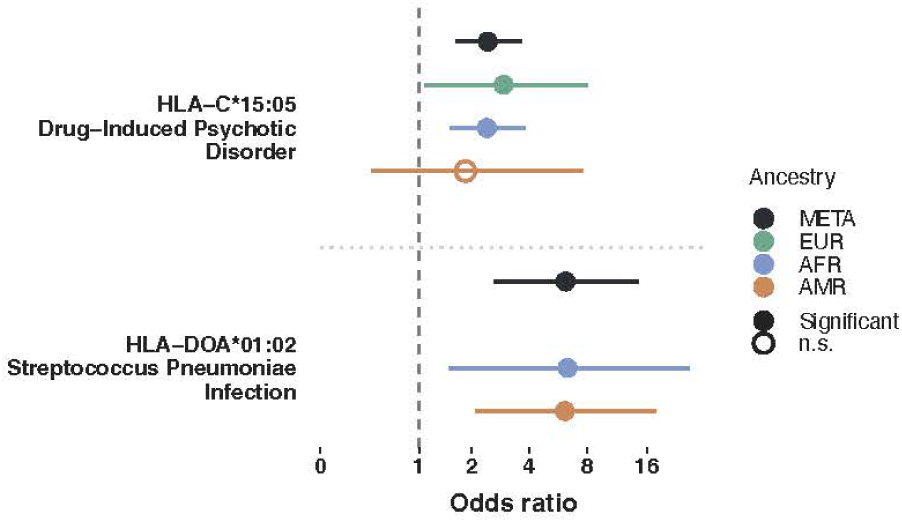
Representative novel HLA–phenotype associations across ancestry groups. Forest plots show odds ratios and 95% confidence intervals for two novel HLA allele-phenotype associations identified in the cross-ancestry meta-analysis. Estimates are shown for the meta-analysis (META) and separately for the EUR, AFR, and AMR ancestry groups. The dashed vertical line marks the null effect (OR = 1). Filled circles indicate associations significant at FDR q < 0.05; open circles indicate non-significant associations.

## DISCUSSION

The results of a high-resolution, multi-ancestry HLA analysis conducted in a large United States-based biobank reveal insights with the potential to change interpretation of MHC association patterns. Using WGS rather than SNP-based HLA imputation, we captured rare and low-frequency alleles, interrogated classical and non-classical loci, and compared HLA associations across ancestries using a common sequence-based framework. The analysis combined sequence-based HLA inference, broad, phenome-wide analysis, and explicit cross-ancestry comparison in a diverse cohort to extend prior HLA PheWAS work from Karnes et al.^35^ and large-scale studies in the UK Biobank, FinnGen, and Taiwan Biobank ^22,36–38^. Importantly, the analysis results indicate that much of the apparent heterogeneity in HLA-disease association across populations may be explained by differences in allele frequency, linkage disequilibrium, and statistical power rather than by fundamentally distinct ancestry-specific architectures. This interpretation is consistent with the trajectory of recent HLA studies: early PheWAS showed broad pleiotropy, but studies were limited largely to European-ancestry imputed data^35^, whereas later biobank studies expanded scale or resolution but were more limited in phenotype breadth, ancestry structure, and analytic focus than the present study.^36–39^

Within the detectability framework employed here, shared HLA-mediated disease architecture may appear ancestry-restricted or vary in apparent effect size because of differences in allele frequency, local LD structure, and sample size. However, the incomplete concordance of results across ancestry groups suggests that a subset of associations may reflect genuine heterogeneity across populations, underscoring the value of explicit multi-ancestry analyses rather than assuming complete transferability of HLA effects.

Another major finding is the substantial reduction of the apparent complexity of HLA association signals with conditional modeling. For representative immune-mediated phenotypes, including type 1 diabetes, celiac disease, hypothyroidism, multiple sclerosis, and rheumatoid arthritis, dozens of marginally associated alleles collapsed to a small number of independent signals, largely concentrated in class II loci with additional class I contributions. This finding is consistent with prior fine-mapping work showing that a substantial proportion of HLA risk can often be localized to a limited set of functionally relevant haplotypic or amino acid effects rather than a very large number of truly independent allele effects.^40,41^ Results of our LD analyses, particularly in the class II region spanning *DRB1*, *DQA1*, and *DQB1*, where extended haplotypes generate dense clusters of correlated marginal associations, provide a structural explanation for this pattern. In practical terms, these findings reinforce that single-allele scans in HLA are difficult to interpret mechanistically without multivariable or haplotype-aware modeling.

Our observation of extensive pleiotropy, with most associated alleles linked to multiple phecodes, likely reflects a combination of shared immunologic mechanisms, correlated clinical phenotypes, and extended LD across the MHC rather than wholly independent biological effects for each association. Pleiotropic associations were concentrated in class II loci, which is consistent with the central role of antigen presentation in immune-mediated disease, but also extended into the infectious, endocrine, dermatologic, and neurologic domains. For example, within the interactive PheWAS Explorer, we observed that *HLA-DRB1.03.01.01G*, which is strongly associated with celiac disease, also has an apparent protective association with Clostridioides difficile infection. Because the celiac signal likely reflects the established *DR3-DQ2* background,^42^ whereas the infection association is comparatively novel, this finding is best viewed as hypothesis-generating and illustrative of the broad value of phenome-wide HLA exploration.

The interactive PheWAS Explorer (https://manticore.niehs.nih.gov/AoU_HLA_PheWAS_Explorer) supports queries of HLA-phenotype associations by allele, phecode, and ancestry and enables visualization of ancestry and trait comparisons. The tool allows users to move from broad signal discovery to targeted follow-up within a single interface, facilitating interpretation and reuse of the findings by users. The PheWAS Explorer also provides complementary views of the data, including Manhattan-style summaries, trait-centered forest plots, multi-trait heatmaps, ancestry-specific contrasts, allele frequency exploration, and higher-resolution allele drilldown. Together, these functions allow users to identify the strongest associations, compare effect estimates across ancestry groups, examine whether related alleles show convergent or divergent phenotypic profiles, and prioritize findings for replication or biological follow-up.

Despite the important implications of the findings, the work has several limitations. First, while short-read WGS improves population-scale HLA analysis, it does not fully resolve complex haplotypes, structural variation, gene presence/absence, or long-range phase across the MHC. Second, G-group resolution captures shared antigen-recognition-domain sequence but may obscure potentially relevant variation outside these domains. Third, ancestry representation was uneven in the study, with larger samples and thus greater power in EUR than in AFR or AMR populations. Fourth, the EHR-derived phenotypes used in the study are susceptible to coding variability, correlated ascertainment, and healthcare utilization effects. Finally, while conditional modeling reduced association complexity, it did not directly identify causal amino acid positions or fully resolve haplotypic mechanisms. This work also suggests the need to update the use of PRS, which typically capture HLA risk through tag variants in imputed data, by explicitly including high-resolution HLA alleles inferred from WGS. This inclusion would meaningfully improve risk model calibration and transferability across genetic ancestries.

Overall, the results show that multi-ancestry, sequence-based HLA analysis has more benefits than merely expanding discovery. By separating sampling-driven differences in detectability from true biological heterogeneity and collapsing dense allele-level signals into a more compact cross-ancestry architecture, this study provides a precise framework for understanding how HLA variation shapes human disease.

## Data Availability

The data used in this study are available to approved researchers through the All of Us Researcher Workbench community workspace (https://workbench.researchallofus.org/). Data can also be explored in the PheWAS Explorer (https://manticore.niehs.nih.gov/AoU_HLA_PheWAS_Explorer)

https://manticore.niehs.nih.gov/AoU_HLA_PheWAS_Explorer

## Acknowledgments

The authors wish to thank Hannah Collins Cakar for assistance with writing, editing, and manuscript preparation. This research was supported by the Intramural Research Program of the NIH, National Institute of Environmental Health Sciences. The contributions of the NIH author(s) are considered Works of the United States Government. The findings and conclusions presented in this paper are those of the author(s) and do not necessarily reflect the views of the NIH or the U.S. Department of Health and Human Services. This research was also supported by the National Cancer Institute of the National Institutes of Health under Award Number R15CA293800.

---

**Supplementary Figure 1.**
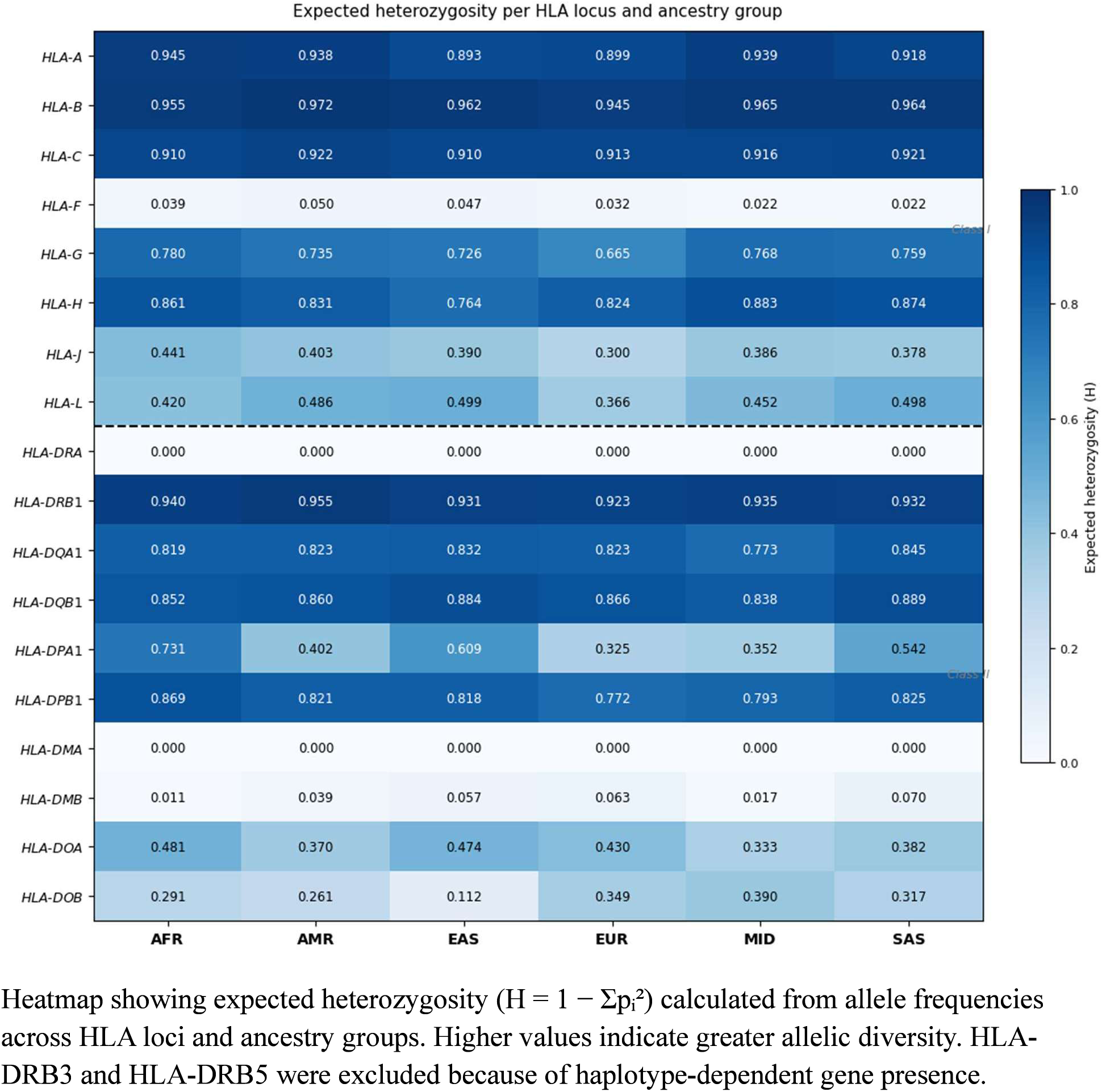
Expected heterozygosity across HLA loci and ancestry groups. Heatmap showing expected heterozygosity (H = 1 - I:p?) calculated from allele frequencies across HLA loci and ancestry groups. Higher values indicate greater allelic diversity. HLA-DRB3 and HLA-DRB5 were excluded because of haplotype-dependent gene presence.

**Supplementary Figure 1.**
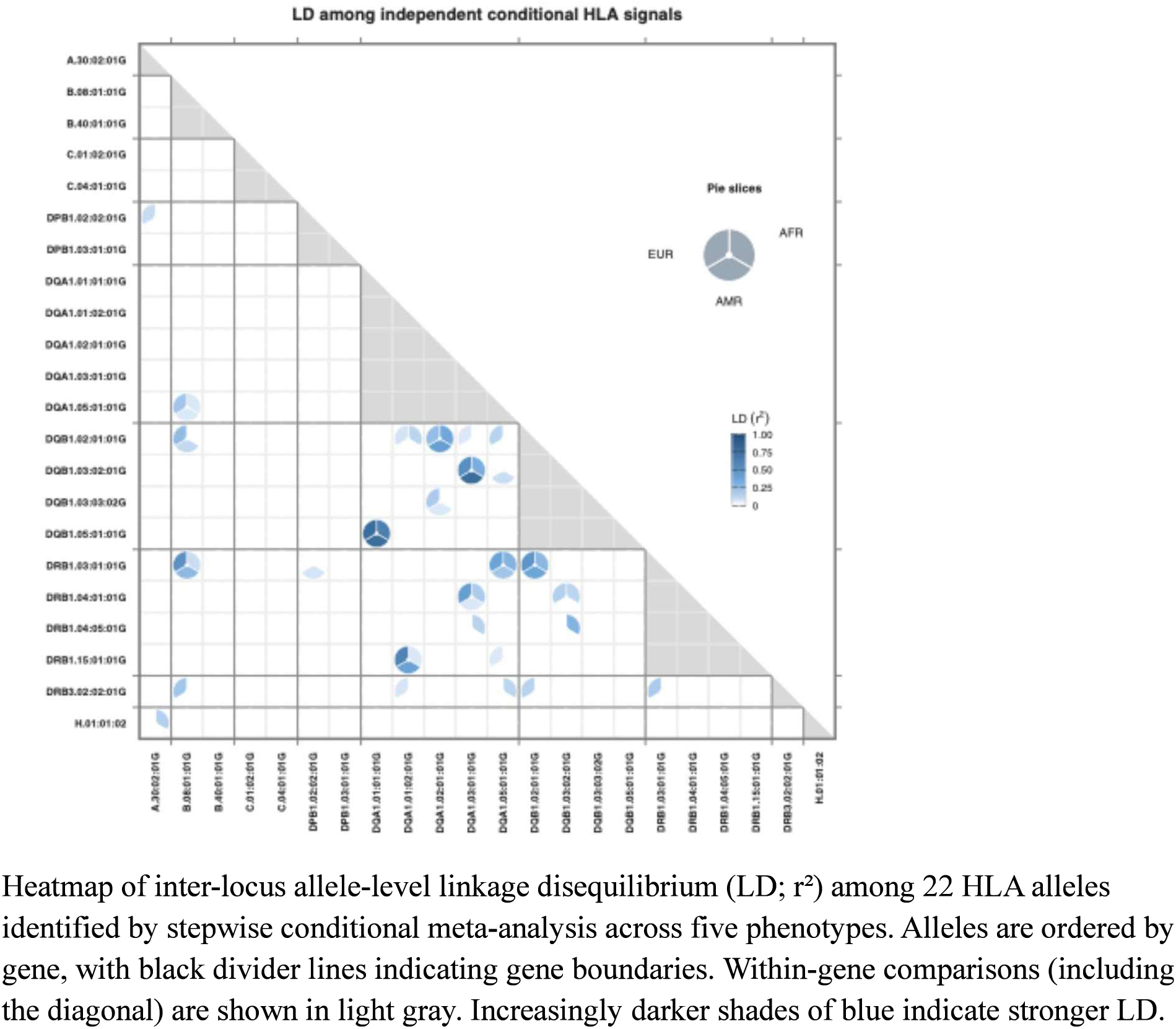
LD among independent conditional HLA alleles across ancestries. Heatmap of inter-locus allele-level linkage disequilibrium (LD; r^2^) among 22 HLA alleles identified by stepwise conditional meta-analysis across five phenotypes. Alleles are ordered by gene, with black divider lines indicating gene boundaries. Within-gene comparisons (including the diagonal) are shown in light gray. Increasingly darker shades of blue indicate stronger LD.

**Supplementary Table 1.** Cohort sizes for WGS analyses after removal of related individuals. Number of unrelated participants included in HLA diversity and linkage disequilibrium analyses after exclusion of related individuals (identity-by-descent> 0.2). AFR, African; AMR, Admixed American; EAS, East Asian; EUR, European; MID, Middle Eastern; SAS, South Asian.

| AFR | AMR | EAS | EUR | MID | SAS | Total |
| --- | --- | --- | --- | --- | --- | --- |
| 77657 | 72151 | 9675 | 224559 | 1458 | 5323 | 390823 |

